# Evaluation of a Mental Health Support Service for Performing Artists

**DOI:** 10.1101/2024.09.21.24314112

**Authors:** Finola M Ryan, Claire Cordeaux, Dermott Davison

**Affiliations:** British Association for Performing Arts Medicine, Cordell Health Ltd, University College London; British Association for Performing Arts Medicine

## Abstract

**Background:** Performing artists frequently face mental health challenges, with risk factors including performance pressure, industry competitiveness, and irregular work patterns. Barriers to seeking help encompass confidentiality concerns, fear of misunderstanding by clinicians, and scheduling conflicts due to peripatetic work.

**Aims:** This study aims to evaluate the accessibility, utilisation, and effectiveness of a Mental Health Support Service (MHSS) for performing artists.

**Methods:** A 12-month evaluation of a UK-wide MHSS for performing artists was conducted. 555 self-referred performers received 6-8 sessions of talking therapy from experienced professionals. Pre- and post-intervention mental health was assessed using PHQ-9 and GAD-7 scales. Quantitative data were analysed using paired t-tests, and qualitative feedback underwent thematic analysis.

**Results:** 240 performers completed pre- and post-intervention scores. Significant reductions in PHQ-9 (mean decrease 6.242, p<0.0001) and GAD-7 (mean decrease 6.225, p<0.0001) scores were observed post-intervention. Qualitative feedback (n=215) revealed high satisfaction, with 88% willing to recommend the service. Key themes included appreciation for tailored support and therapist-performer compatibility. Limited data on outcomes for global majority participants and gender differences necessitate further research.

**Conclusions:** This evaluation of a mental health support service for performing artists reveals high satisfaction rates and willingness to recommend the service, highlighting the value of tailored interventions that address industry-specific challenges. The findings underscore the importance of developing sector-specific mental health standards and support systems, particularly for industries with high proportions of freelance workers, which could have broader implications for improving mental health support across diverse professional fields.

## Introduction

Performing artists face diverse psychological challenges that can impact their mental health(1, 2). These include performance anxiety, exacting standards, and irregular work patterns. Many in the UK performing arts sector are self-employed, or freelance(3), often balancing economic and artistic pressures at the expense of personal wellbeing(4).

A novel mental health support service (MHSS) specifically for performing artists, was launched in January 2020, combining sector-specific expertise with thorough clinical oversight (figure 1). It offers specialised talking therapy delivered by mental health professionals with either personal experience working as a performer, or professional experience working with individuals in that occupational group(5, 6). The MHSS is funded by sector charities and unions. Robust clinical governance is a core element, including triage and case review by a multidisciplinary team (MDT).

**Figure 1.**
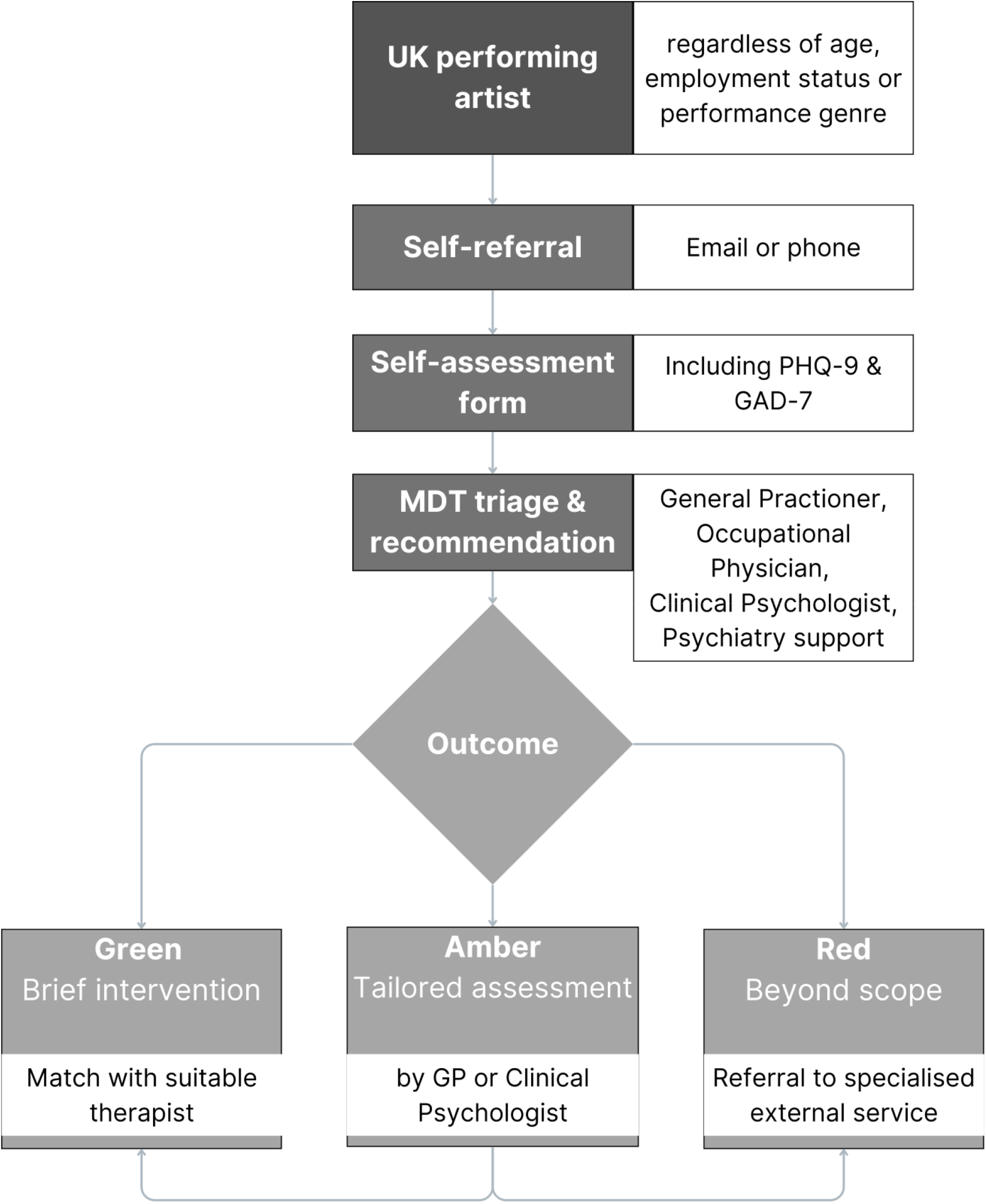
Triage Process Flow Chart for UK Performing Artists MHSS. This diagram illustrates the multi-stage triage system, including self-referral, standardised assessment (PHQ-9 and GAD-7), multidisciplinary team evaluation, and three-tier outcome categorisation (green, amber, red) determining appropriate intervention pathways.

Evidence evaluating the effectiveness of interventions tailored to this occupational population is limited. Therefore, we aim to evaluate the MHSS accessibility, utilisation, and effectiveness for performing artists.

## Methodology

The evaluation examined an MHSS for UK performing artists over 12 months (July 2023 – June 2024). 555 performing artists self-referred, providing demographic information and completing a self-assessment form reviewed by an MDT.

The intervention consisted of 6-8 sessions of in-person or online talking therapy delivered by UK Council for Psychotherapy (UKCP) or British Association for Counselling and Psychotherapy (BACP) registered professionals with relevant experience in the performing arts sector.

The evaluation focused on (1) accessibility and availability, (2) utilisation rates, and (3) quality and effectiveness of the service.

### Data collection included

- Patient Health Questionnaire (PHQ-9) and Generalised Anxiety Disorder (GAD-7) scales administered pre- and post-intervention
- Qualitative feedback gathered via post-intervention questionnaires

Quantitative data were analysed using paired t-tests to assess changes in mental health status. Qualitative data underwent thematic analysis to identify recurring themes related to accessibility, availability, and satisfaction. Statistical analyses were performed using GraphPad Prism Version 10.3.1.

## Results

240 performers completed pre- and post-intervention assessments with 215 providing qualitative feedback. The sample comprised 40% male and 43% female participants, with 17% preferring not to disclose their sex. Age distribution showed 54% in the 25-34 years category, and 25% in the 35-49 years range. The majority of participants self-identified as white. Demographic information is provided in Table 1.

**Table 1.**
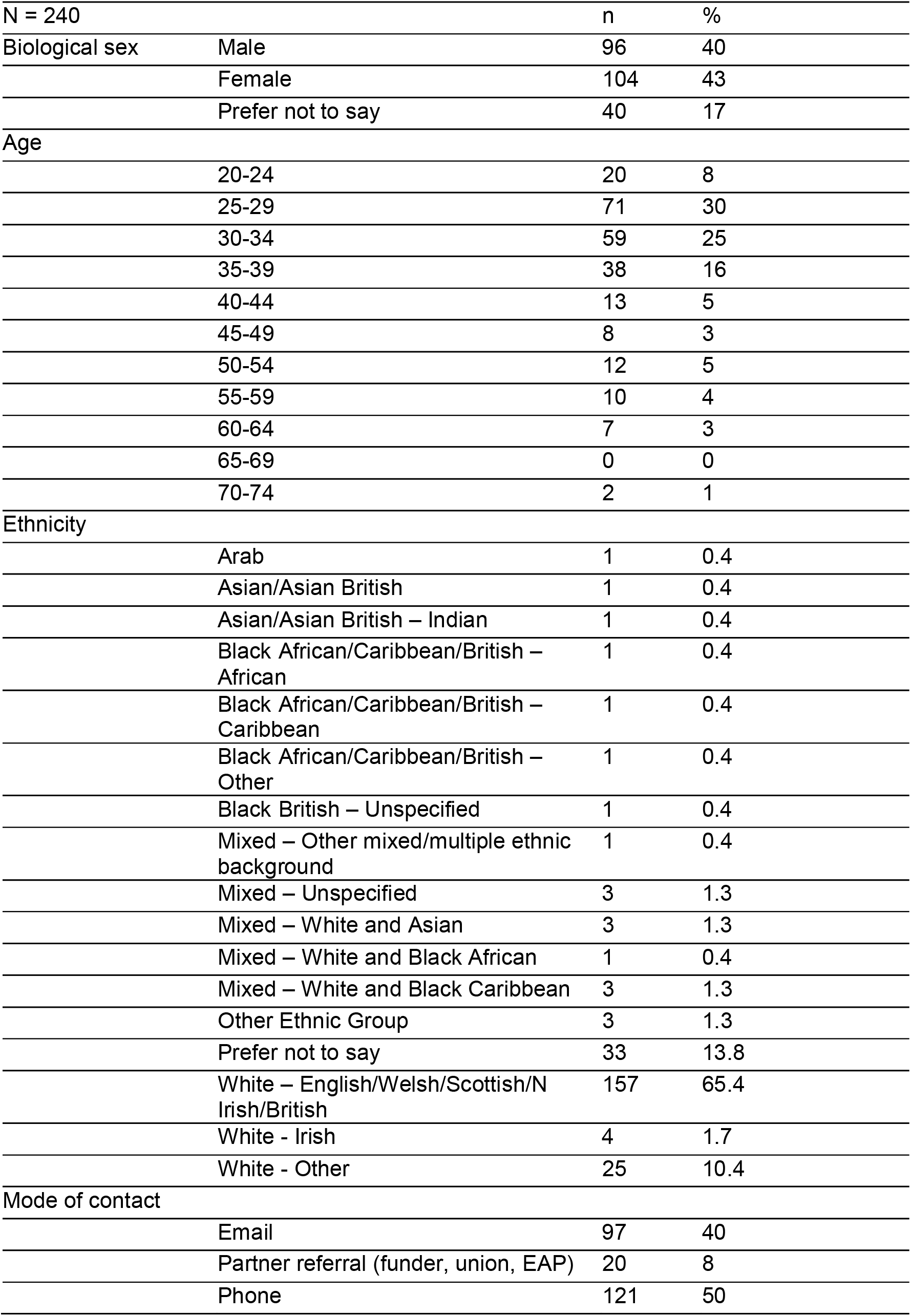
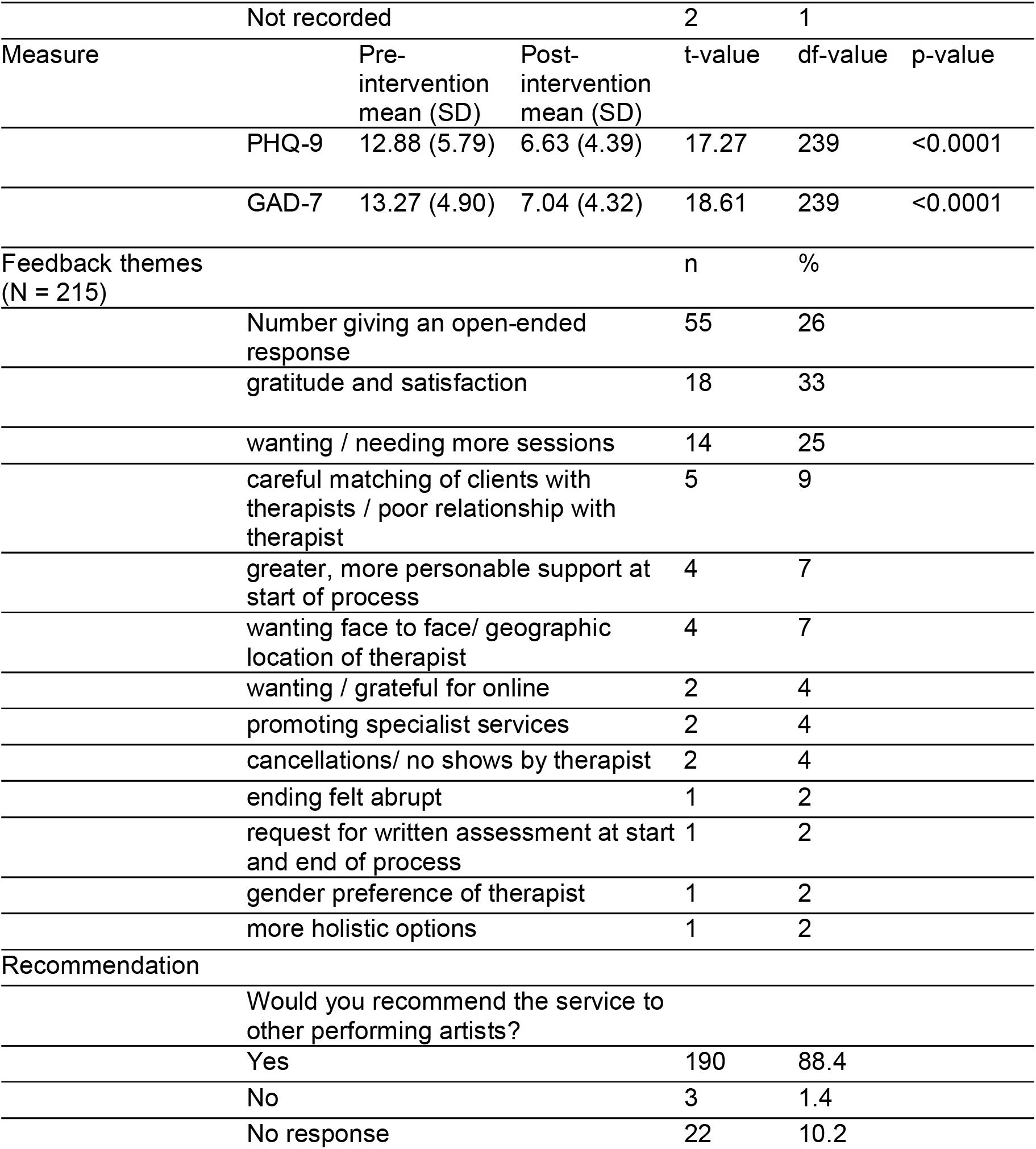
Participant Demographics, Pre- and Post-Intervention Analysis, and Qualitative Feedback.

|  | n | % |
| --- | --- | --- |
| Biological sex |  |  |
| Male | 96 | 40 |
| Female | 104 | 43 |
| Prefer not to say | 40 | 17 |
| Age |  |  |
| 20-24 | 20 | 8 |
| 25-29 | 71 | 30 |
| 30-34 | 59 | 25 |
| 35-39 | 38 | 16 |
| 40-44 | 13 | 5 |
| 45-49 | 8 | 3 |
| 50-54 | 12 | 5 |
| 55-59 | 10 | 4 |
| 60-64 | 7 | 3 |
| 65-69 | 0 | 0 |
| 70-74 | 2 | 1 |
| Ethnicity |  |  |
| Arab | 1 | 0.4 |
| Asian/Asian British | 1 | 0.4 |
| Asian/Asian British – Indian | 1 | 0.4 |
| Black African/Caribbean/British – African | 1 | 0.4 |
| Black African/Caribbean/British – Caribbean | 1 | 0.4 |
| Black African/Caribbean/British – Other | 1 | 0.4 |
| Black British – Unspecified | 1 | 0.4 |
| Mixed – Other mixed/multiple ethnic background | 1 | 0.4 |
| Mixed – Unspecified | 3 | 1.3 |
| Mixed – White and Asian | 3 | 1.3 |
| Mixed – White and Black African | 1 | 0.4 |
| Mixed – White and Black Caribbean | 3 | 1.3 |
| Other Ethnic Group | 3 | 1.3 |
| Prefer not to say | 33 | 13.8 |
| White – English/Welsh/Scottish/N Irish/British | 157 | 65.4 |
| White - Irish | 4 | 1.7 |
| White - Other | 25 | 10.4 |
| Mode of contact |  |  |
| Email | 97 | 40 |
| Partner referral (funder, union, EAP) | 20 | 8 |
| Phone | 121 | 50 |

|  |  |  |  |  |  |  |
| --- | --- | --- | --- | --- | --- | --- |
|  | Not recorded |  |  | 2 | 1 |  |
| Measure |  | Pre-intervention mean (SD) | Post-intervention mean (SD) | t-value | df-value | p-value |
|  | PHQ-9 | 12.88 (5.79) | 6.63 (4.39) | 17.27 | 239 | <0.0001 |
|  | GAD-7 | 13.27 (4.90) | 7.04 (4.32) | 18.61 | 239 | <0.0001 |
| Feedback themes (N = 215) |  |  |  | n | % |  |
|  | Number giving an open-ended response |  |  | 55 | 26 |  |
|  | gratitude and satisfaction |  |  | 18 | 33 |  |
|  | wanting / needing more sessions |  |  | 14 | 25 |  |
|  | careful matching of clients with therapists / poor relationship with therapist |  |  | 5 | 9 |  |
|  | greater, more personable support at start of process |  |  | 4 | 7 |  |
|  | wanting face to face/ geographic location of therapist |  |  | 4 | 7 |  |
|  | wanting / grateful for online |  |  | 2 | 4 |  |
|  | promoting specialist services |  |  | 2 | 4 |  |
|  | cancellations/ no shows by therapist |  |  | 2 | 4 |  |
|  | ending felt abrupt |  |  | 1 | 2 |  |
|  | request for written assessment at start and end of process |  |  | 1 | 2 |  |
|  | gender preference of therapist |  |  | 1 | 2 |  |
|  | more holistic options |  |  | 1 | 2 |  |
| Recommendation |  |  |  |  |  |  |
|  | Would you recommend the service to other performing artists? |  |  |  |  |  |
|  | Yes |  |  | 190 | 88.4 |  |
|  | No |  |  | 3 | 1.4 |  |
|  | No response |  |  | 22 | 10.2 |  |

The MHSS demonstrated high accessibility with 95% of performers receiving a response within one day of contact. Commencement timeframes varied: 80% commenced within a timeframe they considered acceptable. 36% began within 1-2 weeks, 33% within 3-4 weeks and 20% after 4 weeks.

Analysis of PHQ-9 and GAD-7 scores (table 1) revealed statistically significant improvements post-intervention (p<0.0001 for both measures). Mean PHQ-9 scores decreased by 6.242 (SD 5.598; CI [-6.954 - -5.530] while GAD-7 scores reduced by 6.225 (SD 5.183; CI [-6.884 - -5.566] after the intervention, indicating reduced depression and anxiety symptoms.

Qualitative feedback highlighted high levels of satisfaction with the service with 88% of respondents willing to recommend the service. Key themes, summarised in table 1, included appreciation for the tailored approach performers’ needs, tension with limited resources, and the importance of therapist-performer compatibility.

## Discussion

This evaluation of a MHSS for performing artists reveals positive impacts on performers’ mental health, as evidenced by improved GAD-7 and PHQ-9 scores. Qualitative feedback highlighted high satisfaction levels, with 88% recommending the service to peers. This suggests that tailoring a MHSS to specific occupational needs can enhance engagement and outcomes. Occupation specific interventions have been considered useful in other occupations with unique and complex demands, including journalists, health and social care workers and veterinary professionals(7-9).

The service’s accessibility, including various contact methods and online sessions was crucial for this freelance, peripatetic workforce with irregular schedules(10, 11).

While utilisation rates were encouraging among those who completed pre-and post-intervention PHQ-9 and GAD-7 scores, an examination of ethnicity and gender demographics revealed an underrepresentation of men and performers from global majority backgrounds. This discrepancy merits further investigation and action, considering the UK Music Diversity Report’s findings on ethnic diversity in the sector(12).

UK legislation on workplace mental health applies to freelance performers, but its implementation is challenging due to their peripatetic work. In practice, risk assessment defaults to the individual worker. This underscores the need for industry-wide standards to ensure comprehensive support regardless of employment status, similar to sector-specific standards developed for other industries(13).

Like other competitive sectors, there is often a culture of secrecy around health matters(14) and performers may worry about confidentiality breaches that could lead to lost opportunities(15-17). A tailored MHSS can help overcome this by offering robust confidentiality protocols, sector-specific expertise and flexible, discreet access options, fostering trust and normalising mental health support. This evaluation has identified the need to streamline service pathways. Next steps involve implementing comprehensive evidence-based self-care techniques and initiatives emphasising proactive health measures and early intervention strategies.

Future research should examine therapists’ experiences in providing flexible support to performers, and should investigate the support needs of non-clinical staff such as helpline operators and administrators. This holistic approach could optimise the entire support chain, ensuring all workers are sufficiently supported while improving service delivery for performing artists.

This service evaluation has several limitations. As a non-research study, it lacks the rigorous controls of formal research protocols. The absence of a control group limits causal inferences about the intervention’s effects. Self-reported measures may introduce bias, and the short evaluation period does not assess the longevity of improvements. Additionally, missing data is a significant limitation, as not every performer who received the intervention completed both pre- and post-intervention scores, potentially leading to an incomplete representation of outcomes. Future plans include an ethics application for longer-term follow-up to address these limitations and provide more comprehensive insights into demographics.

Despite these limitations, this evaluation underscores the potential of a tailored MHSS for performing artists. The positive outcomes observed, coupled with high levels of satisfaction and recommendation to peers, support the continued provision and potential expansion of such services.

### Key learning points

#### What is already known about this subject

- Performing artists often face mental health issues impacting their personal and professional lives across disciplines and career stages.
- Risk factors include pressure to maintain high standards, competitive industry, irregular work patterns, and performance anxiety.
- Barriers include confidentiality concerns leading to lack of work, fear of misunderstanding by clinicians, and inflexible appointments due to peripatetic work.

#### What this study adds

- This study presents the first evaluation of a Mental Health Support Service (MHSS) specifically tailored for performing artists.
- The evaluation reveals high levels of satisfaction with the service among performing artists who utilised it. A significant proportion of service users indicated they would recommend the MHSS to other performers, suggesting its value and appropriateness.
- The study identifies key factors contributing to the service’s success, including specialised knowledge of performers’ challenges, flexible appointment scheduling, and strict confidentiality measures. These findings can inform the development of similar services in other sectors.

#### What impact this may have on practice or policy

- This study demonstrates the value of developing sector-specific mental health standards, particularly for industries with high proportions of freelance workers. Such standards could improve risk assessment processes and ensure comprehensive support regardless of employment status, benefiting various creative and gig economy sectors.
- The findings support the provision and expansion of tailored mental health services in specialised industries. This could inspire similar initiatives in other sectors with unique occupational stressors, leading to improved mental health support across diverse professional fields.
- Establishing a robust chain of clinical support in specialised mental health services is important. This model ensures safety through appropriate escalation pathways and comprehensive care, potentially serving as a blueprint for implementing similar safety-focused systems in other high-stress industries.

## Data Availability

Data not available: participant consent

## Acknowledgements

We extend our sincere gratitude to the governance structure that provided invaluable oversight and guidance throughout this project: the Board of Trustees of BAPAM (British Association for Performing Arts Medicine), the Medical Committee and Mental Health External Scrutiny Group of BAPAM.

We used Claude 3.5 Sonnet (Anthropic, August 2024) for manuscript editing and flow diagram generation under human supervision. Authors reviewed all AI-generated content and are responsible for the final manuscript.

